# The Association of Cerebral Embolic Protection during Transcatheter Aortic Valve Replacement with Periprocedural Neurological Outcomes

**DOI:** 10.64898/2026.03.11.26348198

**Authors:** Funda Baysal, Kira Osipenko, Severin Laengle, Irene Steiner, Paul Werner, Philipp Bartko, Daniel Zimpfer, Martin Andreas, Iuliana Coti

## Abstract

**Objective:** Cerebral Embolic Protection Devices (CEPs) have been designed to minimize the risk of periprocedural stroke. The clinical significance of these devices, however, is still under debate.

**Aims:** We aimed to compare periprocedural neurological outcomes and mortality in patients undergoing transfemoral transcatheter aortic valve replacement (TF-TAVR) with versus without CEP.

**Methods:** A single-center retrospective analysis of 1101 patients undergoing transfemoral TAVR from August 2017 to May 2025 was performed. CEPs were used routinely at our institution whenever feasible beginning with October 2019. The primary outcome was defined as the incidence of ischemic stroke occurring within 3 days postoperatively. Secondary endpoints included overall neurological outcomes defined as a combined endpoint of stroke, transient ischemic attack (TIA) and delirium within 3 days and short-term all-cause mortality.

**Results:** Overall, 809 underwent TF-TAVR with CEP, while 292 were treated without. The primary endpoint of clinical ischemic stroke occurred less frequently in the CEP group (1.4% vs. 4.1%), and the group difference revealed a significant result in a univariable Cox regression analysis (p = 0.007). No clinically relevant differences were observed in the incidence of TIA (0.5% vs. 0.7%, p = 0.71) and postprocedural delirium (1.6% vs. 2.4%, p = 0.39). Although the 30-day mortality rate in the control group was higher, the difference did not reach statistical significance (3.9% vs. 1.9%, p = 0.06).

**Conclusions:** The use of neuroprotection during TAVR was associated with a reduced early periprocedural ischemic stroke.

## Introduction

Stroke remains one of the most feared complications after TAVR. Recent technological advances have therefore increasingly focused on strategies to prevent thromboembolic events. Understanding the mechanism of cerebrovascular complications following TAVR is essential for developing effective preventive strategies ^1^. Early postprocedural strokes are predominantly ischemic and might be driven by procedural factors such as catheter or valve manipulation, rapid pacing, balloon dilatation and mechanical interaction with the aorta - which can dislodge debris into the cerebral circulation ^2^.

CEPs were developed to reduce the risk of periprocedural stroke by capturing embolic material. Imaging based randomized clinical trials ^3–7^ have demonstrated the feasibility and safety of routine CEP use during TAVR, while mitigating new lesion formation on magnetic resonance imaging (MRI).

Despite these encouraging findings, substantial controversy persists regarding the overall effectiveness of CEPs. While imaging data and several observational cohort studies support routine use, the two most recent large scaled randomized clinical trials have failed to show a significant reduction in clinical apparent stroke. In the PROTECTED TAVR trial ^8^, which randomized 3,000 patients, the Sentinel device (Boston Scientific) did not significantly reduce stroke within 72 hours. Consistent with this, the BHF PROTECT-TAVI ^9^, enrolling approximately 7,000 patients, also reported no difference in overall stroke events and disabling stroke between the CEP and control group.

Given that post-TAVR stroke increases in-hospital mortality nearly threefold, ^10^ and that TAVR is increasingly performed in younger patient populations, further evidence is needed to clarify the clinical impact of neuroprotection devices and to identify the patient subgroups most likely to benefit from CEP.

## Methods

### Study Population

The present study is a retrospective data analysis including patients who underwent TAVR from August 2017 to May 2025 at the Department of Cardiac and Thoracic Aortic Surgery at the Medical University of Vienna. For this analysis we excluded patients treated with TAVR via a non-transfemoral approach, and those with an unsuccessful procedure defined by failure of valve delivery or intraprocedural switch to surgical aortic valve replacement (SAVR).

### Data source

Data on baseline characteristics, operative details, and adverse events have been collected retrospectively within the framework of the ongoing VICTORY-II register (Ethics Number: 1680/2020), which was approved by the ethics committee of the Medical University of Vienna.

At hospital admission patients gave a written informed consent and undergone a baseline clinical fragility assessment. After valve implantation, patients are prospectively followed up at discharge, 30 days, 6 months, 1 year and then yearly up to 10 years.

### Operative technique

The indication for TAVR was determined by an interdisciplinary heart team in accordance with current guidelines. Transfemoral TAVR was performed percutaneously – most commonly via the right femoral artery under conscious sedation and local anaesthesia. Both self-expendable and balloon-expandable devices were used. Pre- and post-dilatation was applied when necessary.

Since October 2019, cerebral embolic protection has been routinely implemented at our center. Aortic arch anatomy and branch vessels morphology were assessed by computed tomography (CT), and neuroprotection was omitted in patients with unsuitable anatomy such as severe stenosis and tortuosity, soft plaques or when vascular access was insufficient.

Two different neuroprotection devices were used. The Sentinel device (Boston Scientific, Marlborough, MA), inserted via the right radial artery which consists of two filter units with 140-µm pores, that are positioned in the brachiocephalic trunk and left common carotid artery. In contrast, the TriGuard 3^TM^ device (Keystone Heart Ltd., Caesarea, IL, USA) is a deflection device inserted via the femoral artery and positioned in the aortic arch to cover all three large branches.

### Definitions and Outcomes

All outcome variables were defined according to the Valve Academic Research Consortium- 3 (VARC 3) criteria. Neurologic events followed the Neurologic Academic Research Consortium (NeuroARC) classification and were categorized as:

- Overt CNS injury (NeuroARC Type 1)
- Covert CNS injury (NeuroARC Type 2)
- Neurological dysfunction without CNS injury (NeuroARC Type 3)

The primary endpoint was clinical ischemic stroke within 3 days after TAVR defined as an acute focal or multifocal neurological deficit consistent with a vascular territory in the brain, spinal cord, or retina (NeuroARC Type1a or 1aH). Patients with new neurological symptoms underwent CT or MRI to confirm central nervous system (CNS) infarction and determine etiology.

Secondary endpoints included overall neurological events within 3 days and short-term all-cause mortality. Neurological dysfunction without CNS injury comprised transient ischemic attack (Type 3a/3aH) and delirium (Type 3b). As no routine postoperative neuroimaging was performed, asymptomatic patients were not further evaluated and covert CNS injuries (Type 2) or so-called silent strokes were not included.

Short-term mortality refers to all-cause death after valve implantation and includes in-hospital mortality and mortality at 30-days.

### Statistical Analysis

Continuous variables have been reported using mean and standard deviation (SD) if approximately normally distributed or median and interquartile range (first quartile Q1; third quartile Q3) if otherwise. For group comparisons, Welch t-tests or Wilcoxon rank sum tests were performed, respectively. Categorical variables have been reported using absolute frequencies and percentages. Differences have been assessed using Fisher’s exact tests.

To evaluate the association of neuroprotection with the primary outcome, a univariable Cox-regression model with neuroprotection as independent variable was calculated. The dependent variable was the number of days from valve implantation (day 0) until the occurrence of ischemic stroke within an observation period of 3 days. Patients without event were censored at the earliest of re-surgery (3 patients needed re-surgery within 2 days post-TAVR), death or day 3 post-surgery, respectively. In patients with an event occurring on the day of valve implantation, the time frame from surgery until event was set to 0.5 days. An estimate for the hazard ratio with 95% confidence intervals (CI), as well as the p-value (H0: hazard ratio is equal to 1) was reported. Graphical visualization was done by Kaplan Meier plots, whereby a log-rank test was calculated to compare the survival curves. The incidence of ischemic stroke by day 3 post-surgery was calculated as 1 minus Kaplan-Meier estimates, additionally 95% confidence intervals are reported. The secondary endpoints overall neurological events within 3 days, TIA within 3 days, and delirium within 3 days were analyzed analogously. Furthermore, the associations of the following baseline variables with the primary endpoint and with overall neurological events within 3 days, respectively, were analyzed by univariable Cox regression models: age, sex, BMI, EuroScore II (log-transformed), STS Score (log-transformed), Porcelain aorta, Agatston Score, diabetes mellitus, insulin dependent diabetes mellitus, peripheral vascular disease, cerebrovascular disease, history of stroke/TIA, atrial fibrillation/ flutter, coronary artery disease, previous myocardial infarction, CABG, PCI, congestive heart failure, LVEF < 50%, chronic kidney disease, creatinine (log-transformed), dialysis, bicuspid aortic valve, oral anticoagulation with NOAC, oral anticoagulation with VKA, single antiplatelet therapy, valve-in-valve procedure, small valve size (≤ 23mm), use of balloon-expandable valve, balloon dilatation before valve implant, balloon dilatation after valve implant, and postoperative new onset atrial fibrillation. In case of complete separation, a Cox regression model with Firth correction was applied (R-function logistf, R-package logistf version 1.26.1).

To evaluate the association of 30-day mortality and neuroprotection, a univariable Cox-regression model was calculated with neuroprotection as independent factor. The dependent variable was the number of days from valve implantation (day 0) until death within the following 30 days. Patients without event were censored at the earliest of re-surgery (4 patients had a re-surgery at two or four days post-TAVR, respectively), last observation or day 30, respectively. Additionally, univariable Cox-regression models were calculated to analyze the following baseline variables: age, sex, BMI, diabetes mellitus, COPD, coronary artery disease, atrial fibrillation/ flutter, chronic kidney disease, dialysis, PVL end of OP, dialysis immediately after TAVR, Euro Score II (log-transformed), STS (log-transformed), creatinine (log-transformed). To analyze the association of neurological outcomes, all neurological events within 3 days and ischemic stroke within 3 days with 30-day mortality, univariable Cox regression models were calculated with the aforementioned variables included as time-dependent covariables in the model. In-hospital mortality was compared between patients with vs. without CEP as described above. In-hospital mortality was calculated as the number of days from valve implantation (day 0) until death. Patients without event were censored at the earlier of discharge (internal or external) or re-surgery (5 patients). Median length of stay (range) was calculated as days from surgery until death, re-surgery or discharge, respectively.

The results of the univariable Cox regression models of the primary and secondary endpoints have been depicted using forest plots.

As we started using CEP in October 2019, we additionally conducted a subset analysis of patients who underwent TAVR between 2019 and 2025. The results are available in the Supplementary Data.

A p-value of ≤ 0.05 was considered as significant in all statistical tests. Due to the exploratory character of the study, no adjustment for multiple testing was conducted. The interpretation of the p-values is exploratory. Statistical analyses were performed using R statistical software version 4.5.1.

## Results

### 3.1 Study Cohort

A total of 1,107 patients underwent TF-TAVR at our department between August 2017 and May 2025. Six patients with unsuccessful valve deployment were excluded. In five patients, annulus rupture occurred during valve implantation resulting in conversion to SAVR, while in one patient the valve implantation became technically challenging, so the procedure had to be aborted.

Baseline characteristics are summarized in Table 1. Overall, age and sex were equally distributed with a mean age of 81 years in both groups and 52.7% male patients. The median EuroScore II and Society of Thoracic Surgeons score (STS) for the overall cohort were 3.2 (IQR 1.9-5.5) and 8.8 (IQR 7.2-11.1), respectively. Baseline operative risk did not differ substantially between both groups, overall indicating an intermediate to high surgical risk cohort.

**Table 1:**
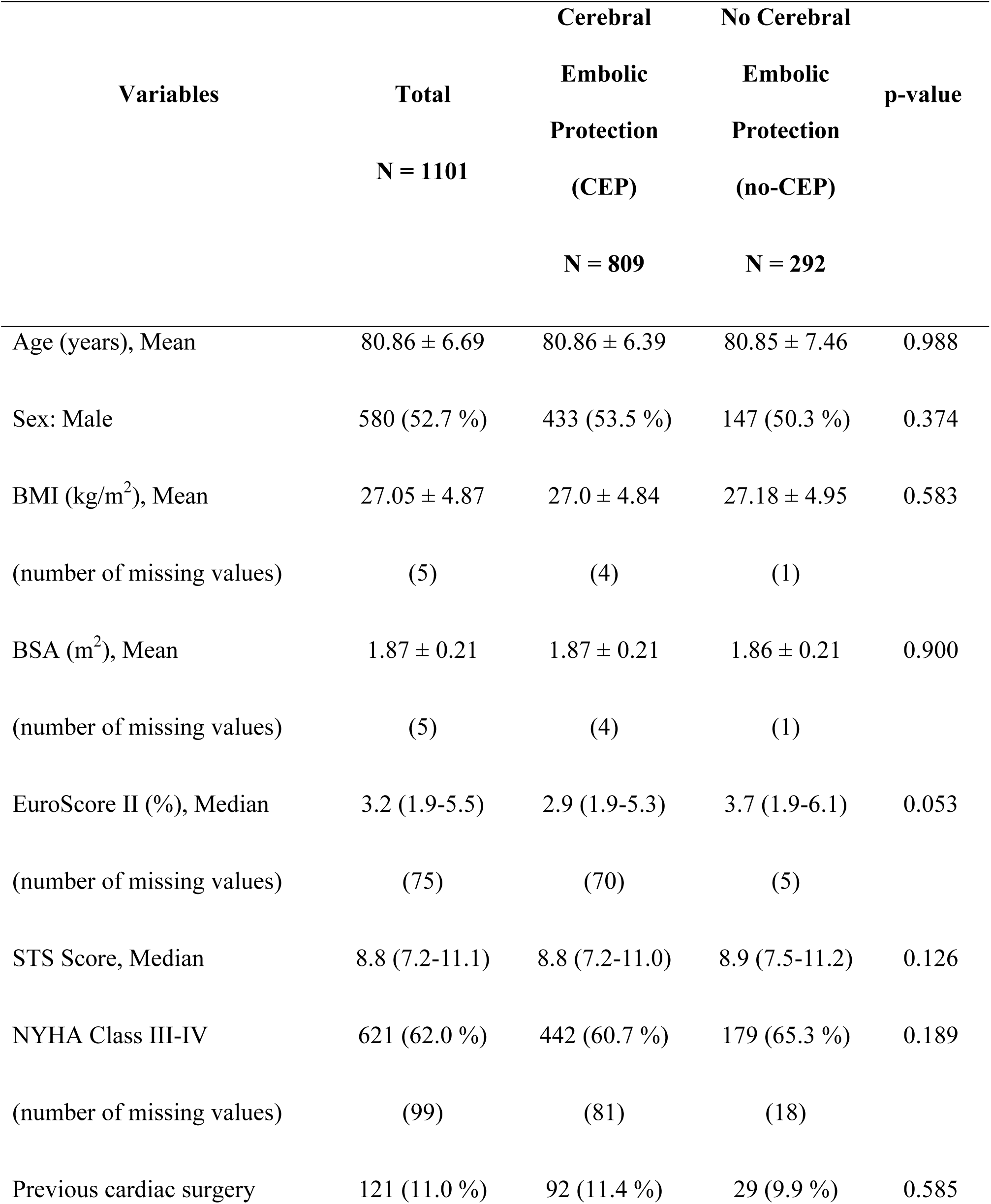
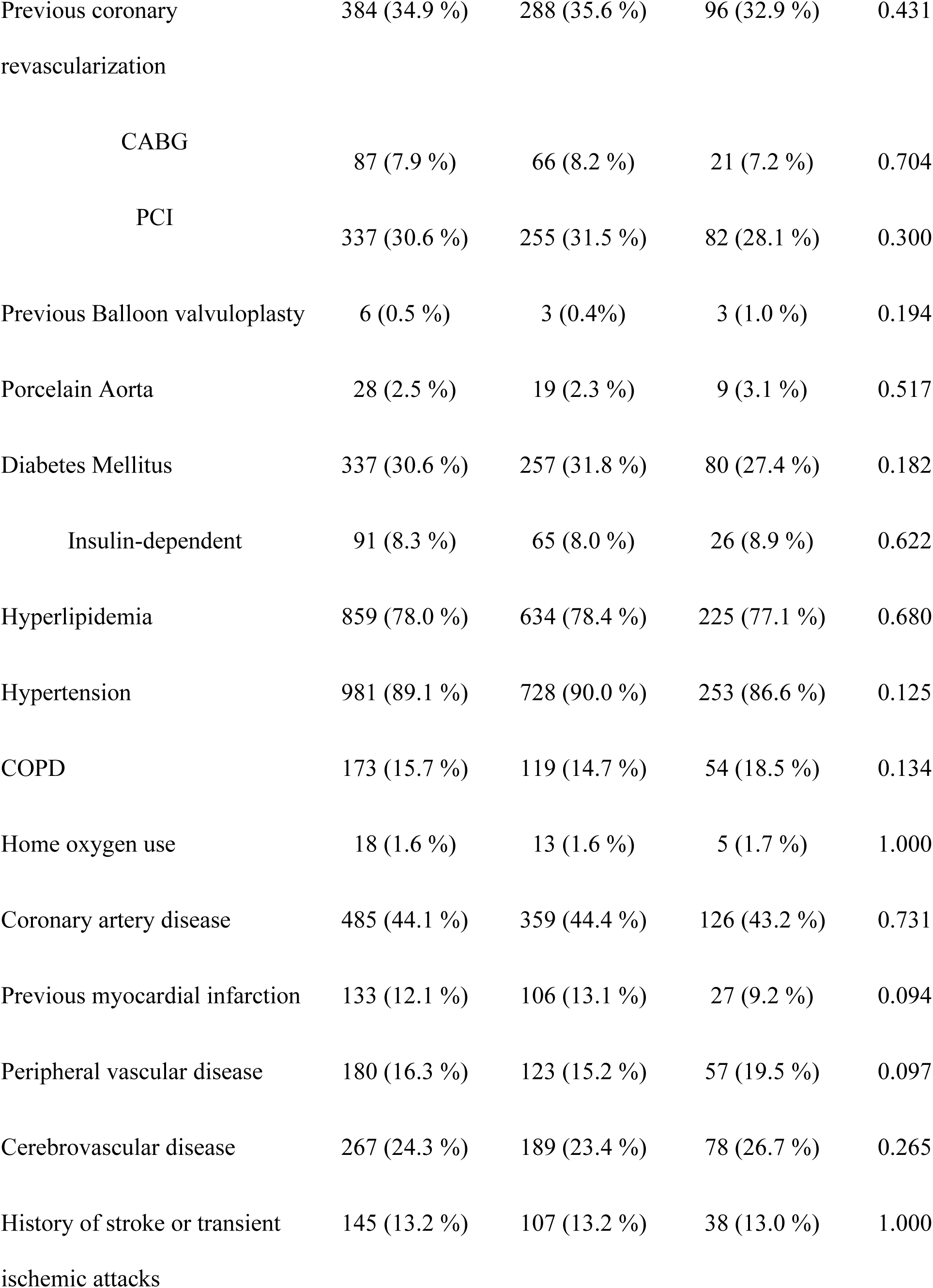
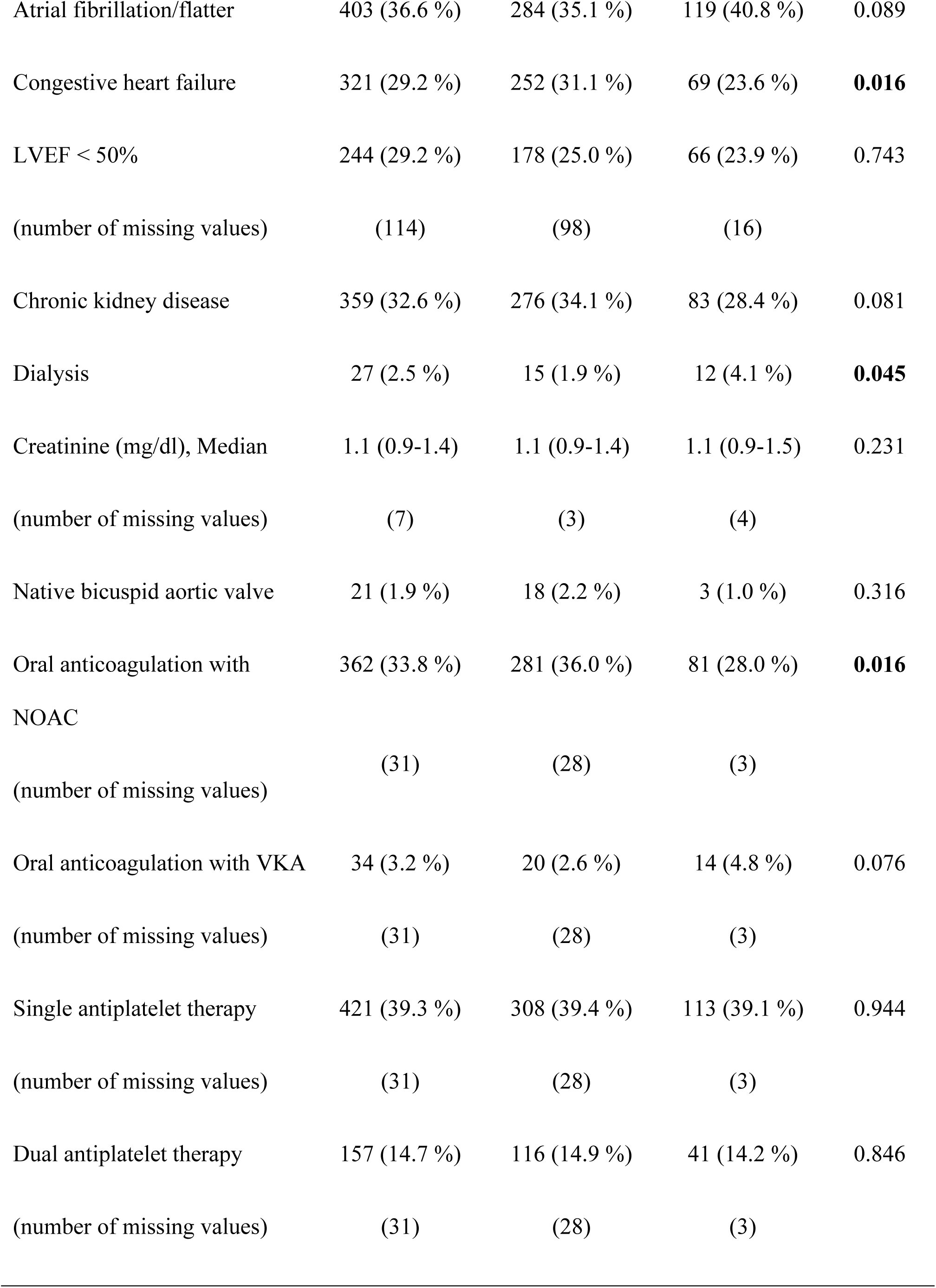
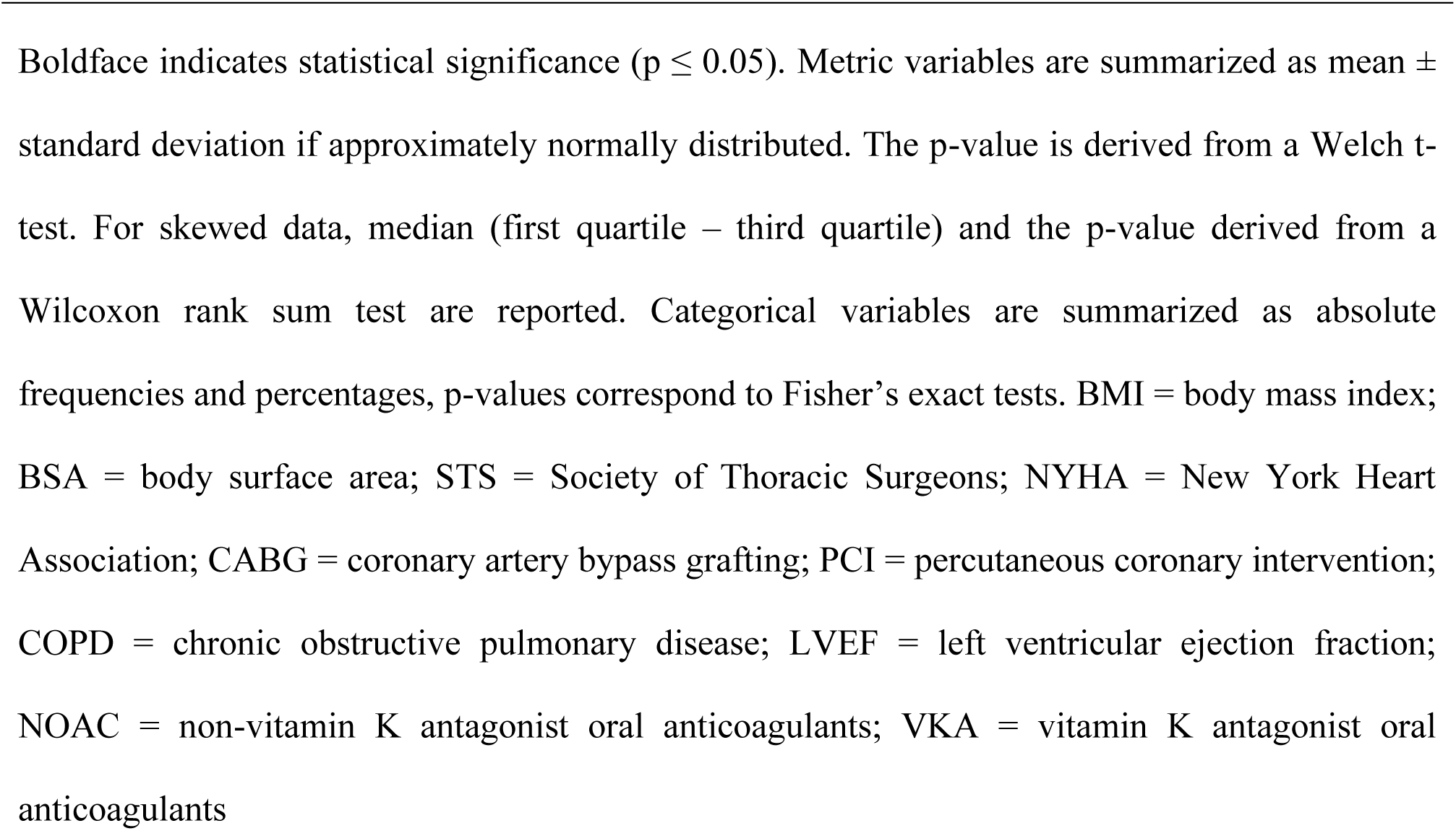
Baseline characteristics.

| Variables | Total<br>N = 1101 | Cerebral<br>Embolic<br>Protection<br>(CEP)<br>N = 809 | No Cerebral<br>Embolic<br>Protection<br>(no-CEP)<br>N = 292 | p-value |
| --- | --- | --- | --- | --- |
| Age (years), Mean | 80.86 ± 6.69 | 80.86 ± 6.39 | 80.85 ± 7.46 | 0.988 |
| Sex: Male | 580 (52.7 %) | 433 (53.5 %) | 147 (50.3 %) | 0.374 |
| BMI (kg/m<sup>2</sup>), Mean | 27.05 ± 4.87 | 27.0 ± 4.84 | 27.18 ± 4.95 | 0.583 |
| (number of missing values) | (5) | (4) | (1) |  |
| BSA (m<sup>2</sup>), Mean | 1.87 ± 0.21 | 1.87 ± 0.21 | 1.86 ± 0.21 | 0.900 |
| (number of missing values) | (5) | (4) | (1) |  |
| EuroScore II (%), Median | 3.2 (1.9-5.5) | 2.9 (1.9-5.3) | 3.7 (1.9-6.1) | 0.053 |
| (number of missing values) | (75) | (70) | (5) |  |
| STS Score, Median | 8.8 (7.2-11.1) | 8.8 (7.2-11.0) | 8.9 (7.5-11.2) | 0.126 |
| NYHA Class III-IV | 621 (62.0 %) | 442 (60.7 %) | 179 (65.3 %) | 0.189 |
| (number of missing values) | (99) | (81) | (18) |  |
| Previous cardiac surgery | 121 (11.0 %) | 92 (11.4 %) | 29 (9.9 %) | 0.585 |
| Previous coronary revascularization | 384 (34.9 %) | 288 (35.6 %) | 96 (32.9 %) | 0.431 |
| CABG | 87 (7.9 %) | 66 (8.2 %) | 21 (7.2 %) | 0.704 |
| PCI | 337 (30.6 %) | 255 (31.5 %) | 82 (28.1 %) | 0.300 |
| Previous Balloon valvuloplasty | 6 (0.5 %) | 3 (0.4%) | 3 (1.0 %) | 0.194 |
| Porcelain Aorta | 28 (2.5 %) | 19 (2.3 %) | 9 (3.1 %) | 0.517 |
| Diabetes Mellitus | 337 (30.6 %) | 257 (31.8 %) | 80 (27.4 %) | 0.182 |
| Insulin-dependent | 91 (8.3 %) | 65 (8.0 %) | 26 (8.9 %) | 0.622 |
| Hyperlipidemia | 859 (78.0 %) | 634 (78.4 %) | 225 (77.1 %) | 0.680 |
| Hypertension | 981 (89.1 %) | 728 (90.0 %) | 253 (86.6 %) | 0.125 |
| COPD | 173 (15.7 %) | 119 (14.7 %) | 54 (18.5 %) | 0.134 |
| Home oxygen use | 18 (1.6 %) | 13 (1.6 %) | 5 (1.7 %) | 1.000 |
| Coronary artery disease | 485 (44.1 %) | 359 (44.4 %) | 126 (43.2 %) | 0.731 |
| Previous myocardial infarction | 133 (12.1 %) | 106 (13.1 %) | 27 (9.2 %) | 0.094 |
| Peripheral vascular disease | 180 (16.3 %) | 123 (15.2 %) | 57 (19.5 %) | 0.097 |
| Cerebrovascular disease | 267 (24.3 %) | 189 (23.4 %) | 78 (26.7 %) | 0.265 |
| History of stroke or transient ischemic attacks | 145 (13.2 %) | 107 (13.2 %) | 38 (13.0 %) | 1.000 |
| Atrial fibrillation/flutter | 403 (36.6 %) | 284 (35.1 %) | 119 (40.8 %) | 0.089 |
| Congestive heart failure | 321 (29.2 %) | 252 (31.1 %) | 69 (23.6 %) | **0.016** |
| LVEF < 50% | 244 (29.2 %) | 178 (25.0 %) | 66 (23.9 %) | 0.743 |
| (number of missing values) | (114) | (98) | (16) |  |
| Chronic kidney disease | 359 (32.6 %) | 276 (34.1 %) | 83 (28.4 %) | 0.081 |
| Dialysis | 27 (2.5 %) | 15 (1.9 %) | 12 (4.1 %) | **0.045** |
| Creatinine (mg/dl), Median | 1.1 (0.9-1.4) | 1.1 (0.9-1.4) | 1.1 (0.9-1.5) | 0.231 |
| (number of missing values) | (7) | (3) | (4) |  |
| Native bicuspid aortic valve | 21 (1.9 %) | 18 (2.2 %) | 3 (1.0 %) | 0.316 |
| Oral anticoagulation with NOAC | 362 (33.8 %) | 281 (36.0 %) | 81 (28.0 %) | **0.016** |
| (number of missing values) | (31) | (28) | (3) |  |
| Oral anticoagulation with VKA | 34 (3.2 %) | 20 (2.6 %) | 14 (4.8 %) | 0.076 |
| (number of missing values) | (31) | (28) | (3) |  |
| Single antiplatelet therapy | 421 (39.3 %) | 308 (39.4 %) | 113 (39.1 %) | 0.944 |
| (number of missing values) | (31) | (28) | (3) |  |
| Dual antiplatelet therapy | 157 (14.7 %) | 116 (14.9 %) | 41 (14.2 %) | 0.846 |
| (number of missing values) | (31) | (28) | (3) |  |

Boldface indicates statistical significance ( $p \leq 0.05$ ). Metric variables are summarized as mean ± standard deviation if approximately normally distributed. The p-value is derived from a Welch t-test. For skewed data, median (first quartile – third quartile) and the p-value derived from a Wilcoxon rank sum test are reported. Categorical variables are summarized as absolute frequencies and percentages, p-values correspond to Fisher’s exact tests. BMI = body mass index; BSA = body surface area; STS = Society of Thoracic Surgeons; NYHA = New York Heart Association; CABG = coronary artery bypass grafting; PCI = percutaneous coronary intervention; COPD = chronic obstructive pulmonary disease; LVEF = left ventricular ejection fraction; NOAC = non-vitamin K antagonist oral anticoagulants; VKA = vitamin K antagonist oral anticoagulants

The proportion of patients with end-stage renal disease on dialysis was higher in the unprotected cohort (4.1% vs 1.9%, p = 0.045), while more patients had congestive heart failure (31.1% vs. 23.6%, p = 0.016) in the CEP group. Preoperative treatment with non-vitamin K oral anticoagulants (NOACs) was more frequent in patients with neuroprotection (36.0% vs 28.0%, p = 0.016).

Overall, 809 (73.48%) patients underwent TF-TAVR with CEP, while 292 (26.52%) were performed without protection. Sentinel device accounted for the vast majority of CEP cases (92.6%), whereas the TriGuard system was used in 7.4%. As illustrated in Figure 1, CEP adoption expanded rapidly after its introduction in October 2019, stabilizing at approximately 95% use between 2021 and 2023. With increasing operator experience and improved identification of anatomical contraindications, CEP use declined modestly in 2024.

**Figure 1:**
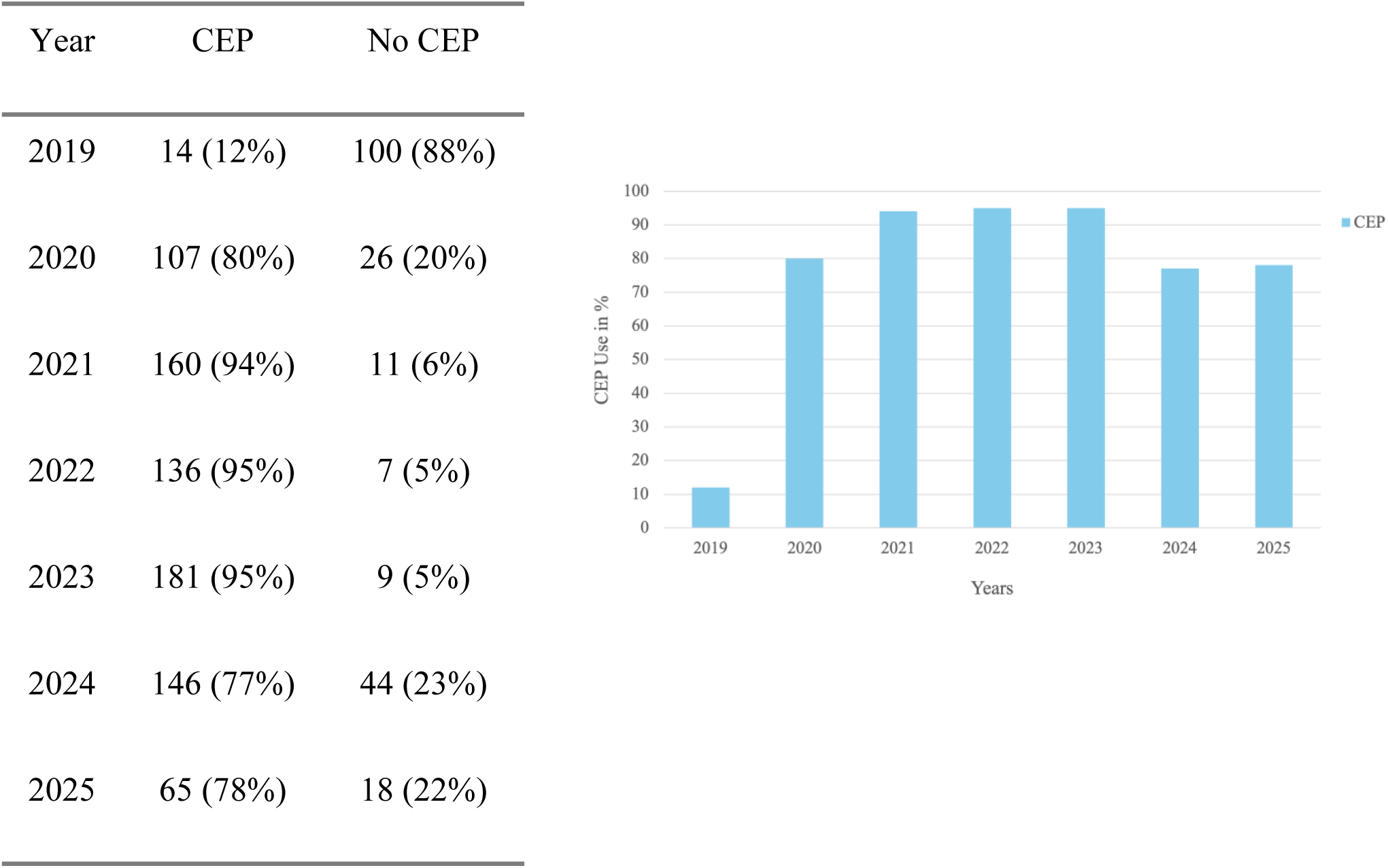
Neuroprotection Use Over Time. CEP use began in October 2019 reaching a plateau of 95% CEP TAVR from 2021 to 2023. CEP = cerebral embolic protection.

Procedural characteristics were similar between groups (Table 2). More than 96% of procedures were not emergent. Valve-in-valve TAVR was performed in 70 patients (6.4%) and pre- and post-implantation balloon dilatation rates were 41.1% and 21.0%, respectively. In 25.1% of patients, a balloon-expandable valve was implanted. Intraoperative paravalvular leakage (≥ moderate) after valve implantation occurred in 16 patients (1.7%).

**Table 2:**
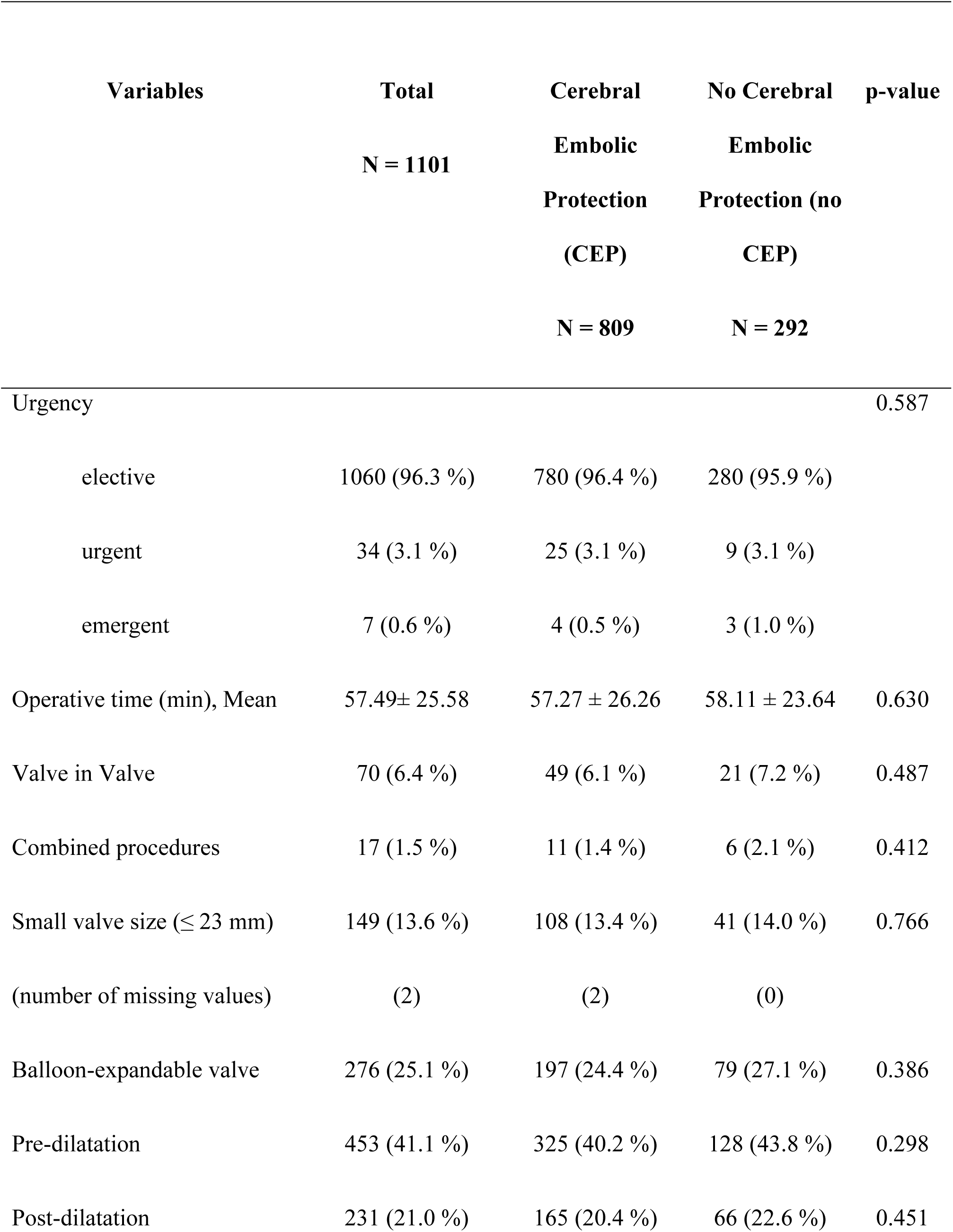
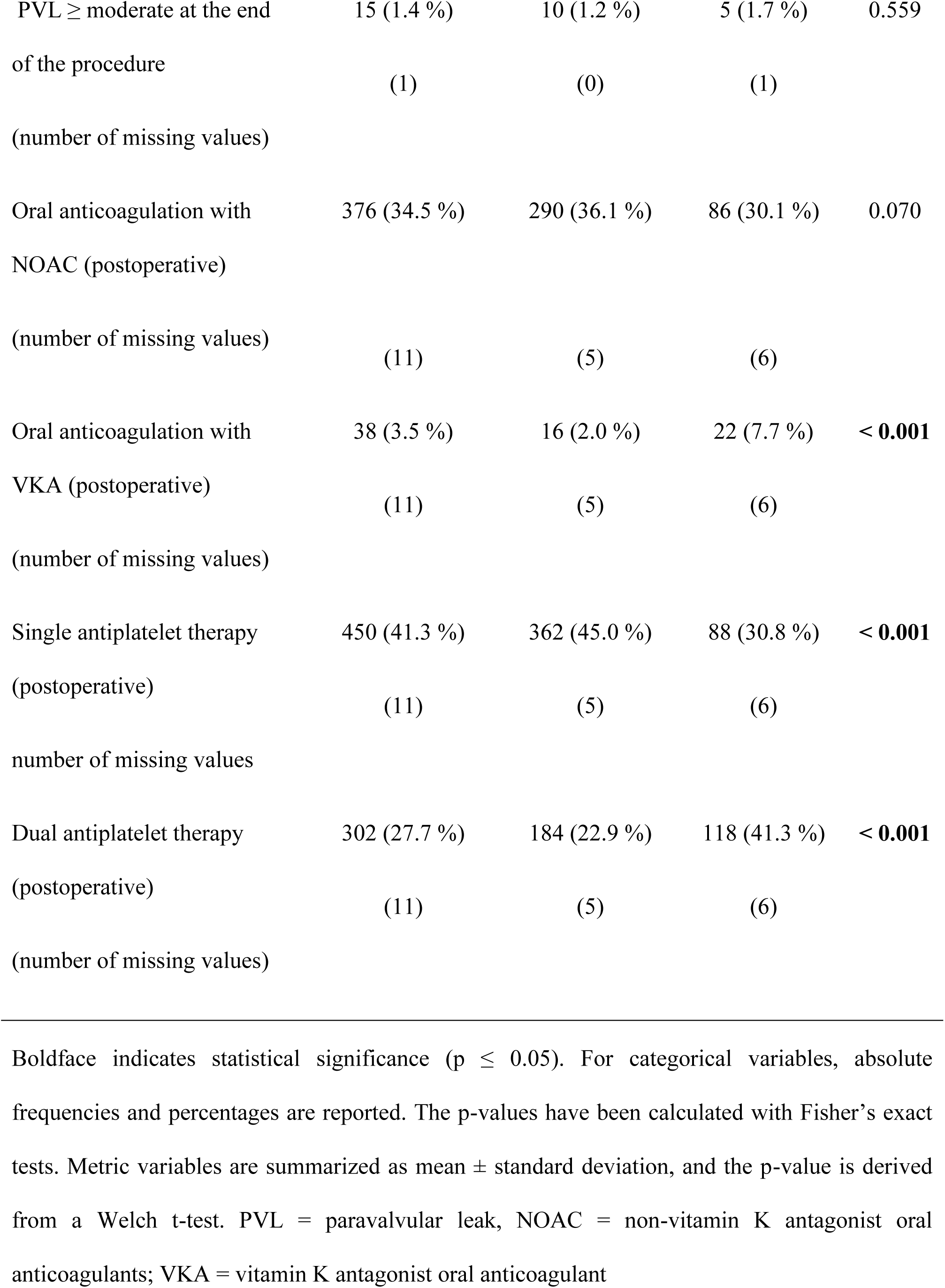
Procedural characteristics.

**Table 3:**
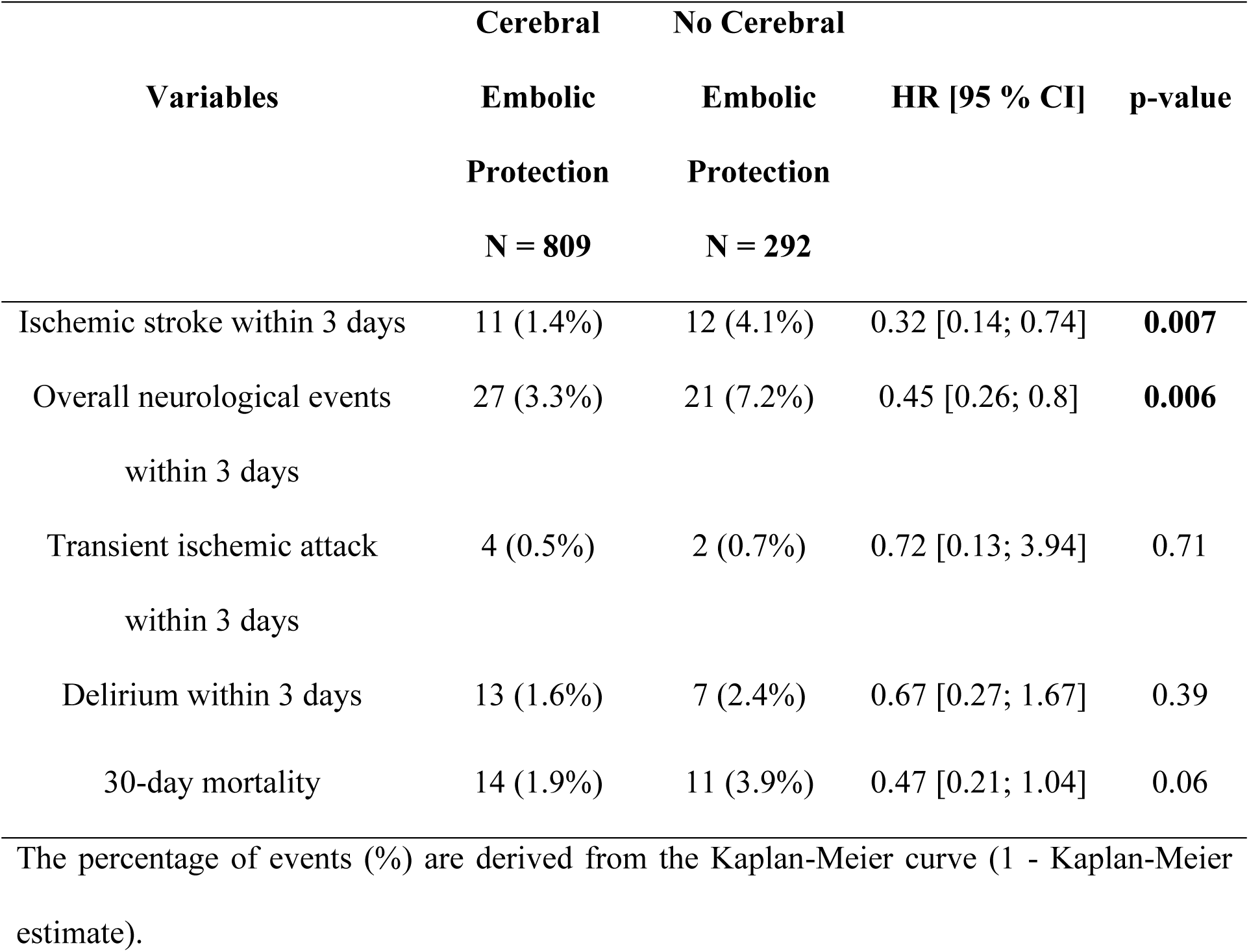
Primary and secondary outcomes.

### 3.2 Primary Outcome – Ischemic stroke

Overall, 23 patients presented with ischemic stroke within 3 days after TAVR. The incidence was markedly higher in the unprotected group, compared with those treated with CEP (4.1% [95% CI: 1.8%; 6.4%] vs. 1.4% [0.6%; 2.2%]). In univariable Cox regression analysis, the use of CEP was associated with a significantly reduced risk of early ischemic stroke [HR 0.32, 95% CI:(0.14-0.74), p = 0.007]. Balloon post-dilatation [HR 2.45, 95% CI:(1.06-5.66), p = 0.04] and a history of cerebrovascular disease [HR 2.42, 95% CI:(1.06-5.52), p = 0.04] were identified as independent predictors of ischemic stroke in the univariable Cox regression analyses (Figure 2).

**Figure 2:**
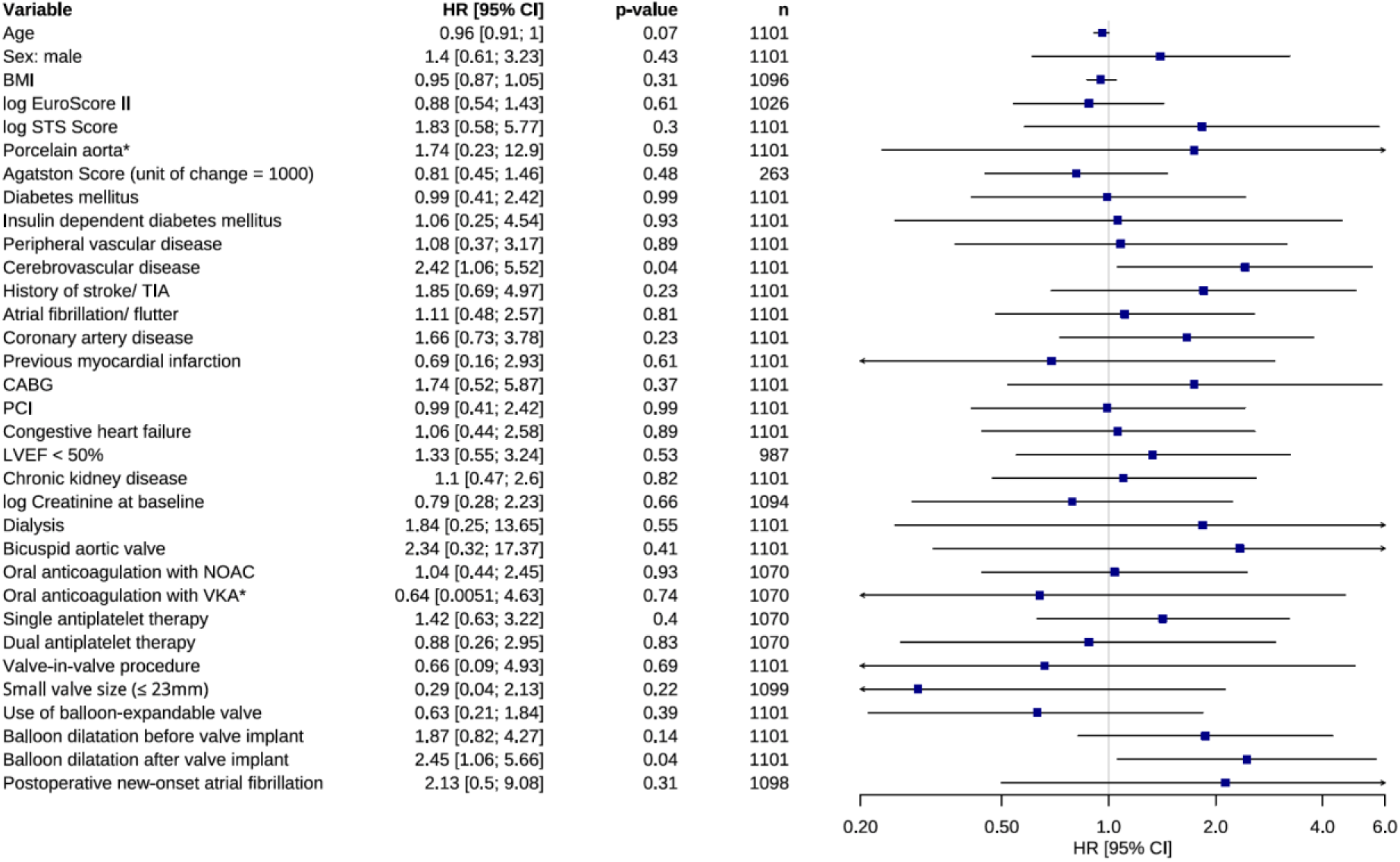
Results of the univariable Cox regression models with ischemic stroke within 3 days after TAVR as dependent variable. Forest plot depicting hazard ratios for ischemic stroke occurring within 3 days after TAVR for prespecified baseline variables. Squares indicate HR and horizontal lines represent 95% CI. P values of the univariable Cox regression models are displayed. BMI = body mass index; STS = Society of Thoracic Surgeons; TIA = transient ischemic attack; CABG = coronary artery bypass grafting; PCI = percutaneous coronary intervention; LVEF = left ventricular ejection fraction; NOAC = non-vitamin K antagonist oral anticoagulants; VKA = vitamin K antagonist oral anticoagulants.

### 3.3 Secondary Outcomes

#### 3.3.1 Overall neurological events

Within 3 days after TAVR, overall neurological events occurred in 3.3% [2.1%; 4.6%] of patients with CEP compared with 7.2% [4.2%; 10.1%] of those without protection. Univariable Cox regression revealed a significantly reduced risk of neurological events in the CEP group [HR 0.45, 95% CI:(0.26-0.80), p = 0.006].

Most neurological events occurred within the first 24 hours. In total, we observed 48 neurological events within 3 days. No hemorrhagic strokes occurred during the 3 days observation period. Further, no significant differences were observed in the incidence of TIA (0.5% [0%; 1%] vs. 0.7% [0%; 1.6%], Cox regression: p = 0.71) and delirium (1.6% [0.7%; 2.5%] vs. 2.4% [0.6%; 4.1%], p = 0.39) between groups.

Univariable analyses (Figure 3) identified cerebrovascular disease [HR 2.27, 95% CI:(1.28-4.04), p = 0.005], history of cerebrovascular event [HR 2.25, 95% CI:(1.17-4.32), p = 0.02] and left-ventricular ejection fraction (LVEF) < 50% [HR 2.15, 95% CI (1.18-3.93), p = 0.01] as predictors of neurological events. Lower body mass index (BMI) also emerged as an unexpected risk factor [HR 0.91, 95% CI:(0.85-0.98), p = 0.01).

**Figure 3:**
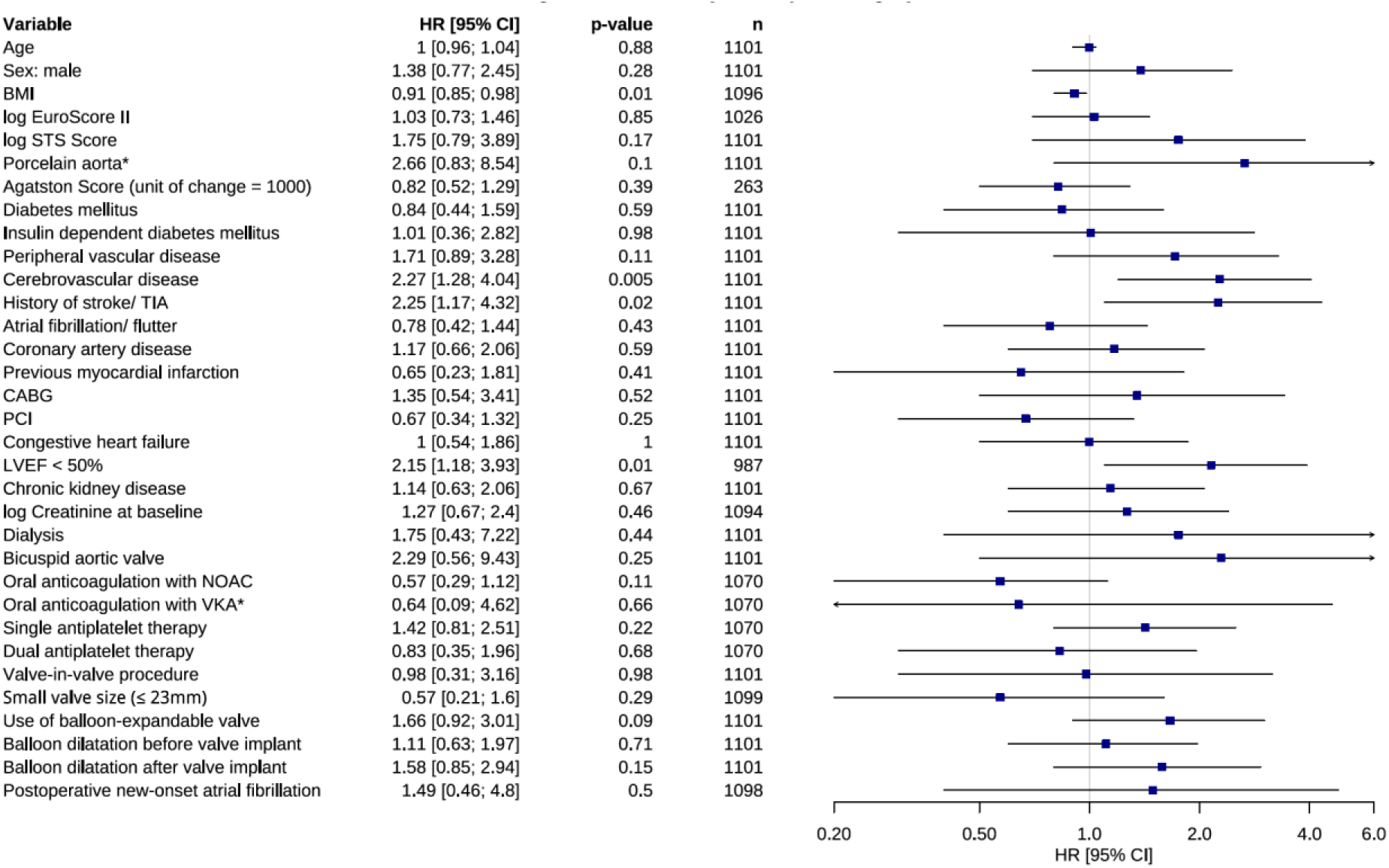
Results of the univariable Cox regression models with overall neurological outcomes within 3 days after TAVR as dependent variable. Forest plot depicting hazard ratios for overall neurological outcomes within 3 days after TAVR for prespecified baseline variables. Squares indicate HR and horizontal lines represent 95% CI. P values of the univariable Cox regression models are displayed.

Since neuroprotection devices were introduced in 2019 at our department, a time-dependent selection bias could not be ruled out. To address this, we conducted a subset analysis of patients undergoing TAVR between 2019 and 2025. The results were concordant with those of the entire study cohort and confirmed a significant negative association of ischemic stroke (p = 0.005) and overall neurological events (p = 0.006) with CEP use (Supplementary Table 1).

#### 3.3.2 Mortality

There was no statistically significant difference in in-hospital mortality between both groups (HR 0.68, 95% CI: (0.3; 1.51), p = 0.34). In total, 24 patients died during hospital stay. The median length of hospital stay was 4 days (range: 1 – 94 days) in the CEP group and 2 days in the control group (range: 1 – 107 days).

The 30-day mortality rate was higher in patients undergoing TAVR without neuroprotection (1.9% [0.9%; 2.9%] with CEP vs. 3.9% [1.6%; 6.2% without CEP]), however, no significant association between the use of neuroprotection and 30-day mortality could be identified [HR 0.47, 95% CI:(0.21-1.04), p = 0.06]. In further univariable Cox regression analyses, higher STS score [HR 7.15, 95% CI:(2.81-18.16), p < 0.0001] and elevated baseline creatinine [HR 2.45, 95% CI:(1.17-5.1), p = 0.02] as well as new-onset renal replacement therapy after TAVR [HR 10.31, 95% CI:(1.39-76.19), p = 0.02] independently predicted 30-day mortality. Ischemic stroke within 3 days was strongly associated with increased 30-day mortality [HR 7.04, 95% CI:(2.11-23.52), p = 0.002] (Figure 4).

**Figure 4:**
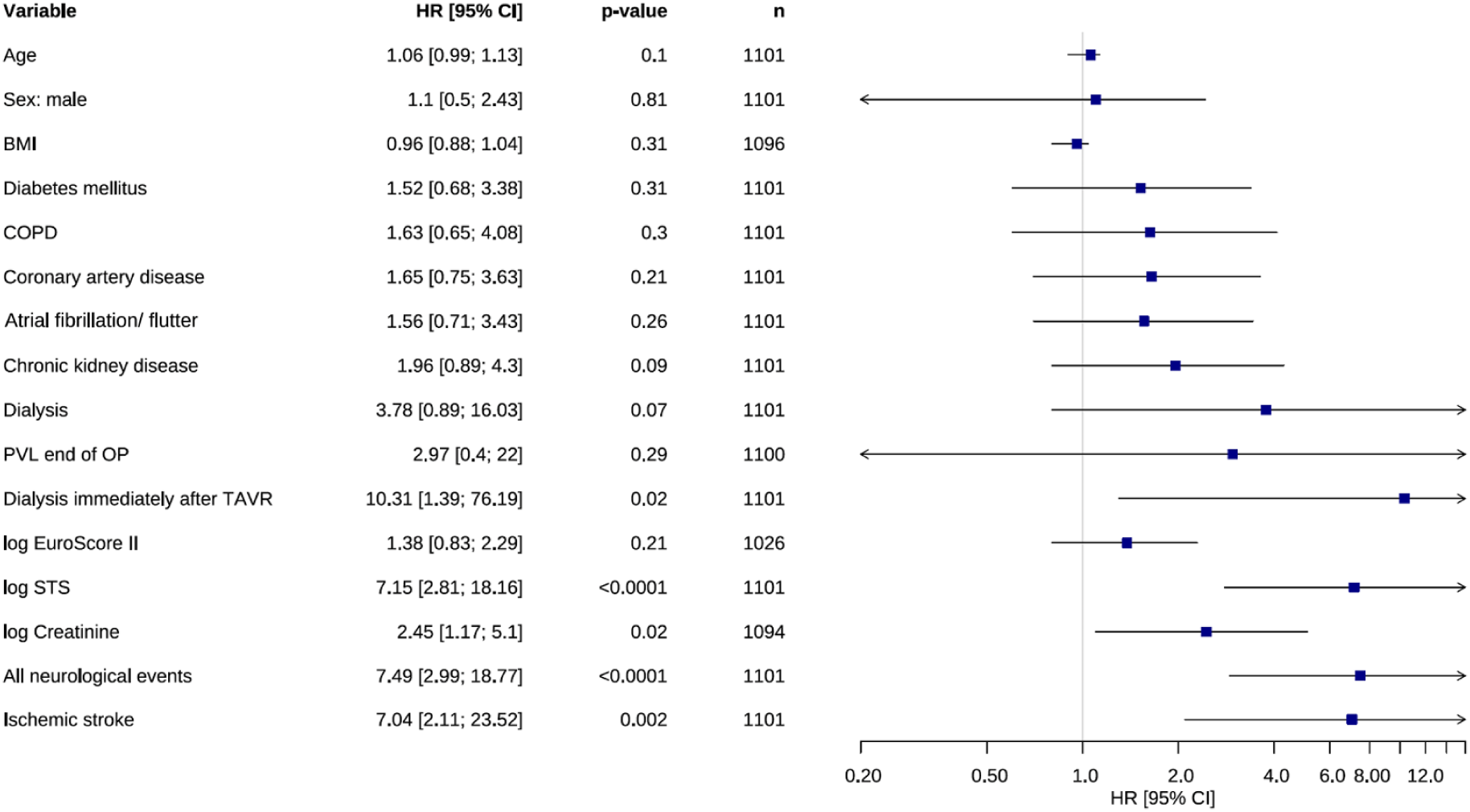
Results of the univariable Cox regression models with 30-day all-cause mortality after TAVR as dependent variable. Forest plot depicting hazard ratios for all-cause mortality within 30 days after TAVR for prespecified baseline variables. Squares indicate HR and horizontal lines represent 95% CI. P values of the univariable Cox regression models are displayed. BMI = body mass index; COPD = chronic obstructive pulmonary disease; PVL = paravalvular leak; STS = Society of Thoracic Surgeons.

**Figure 5:**
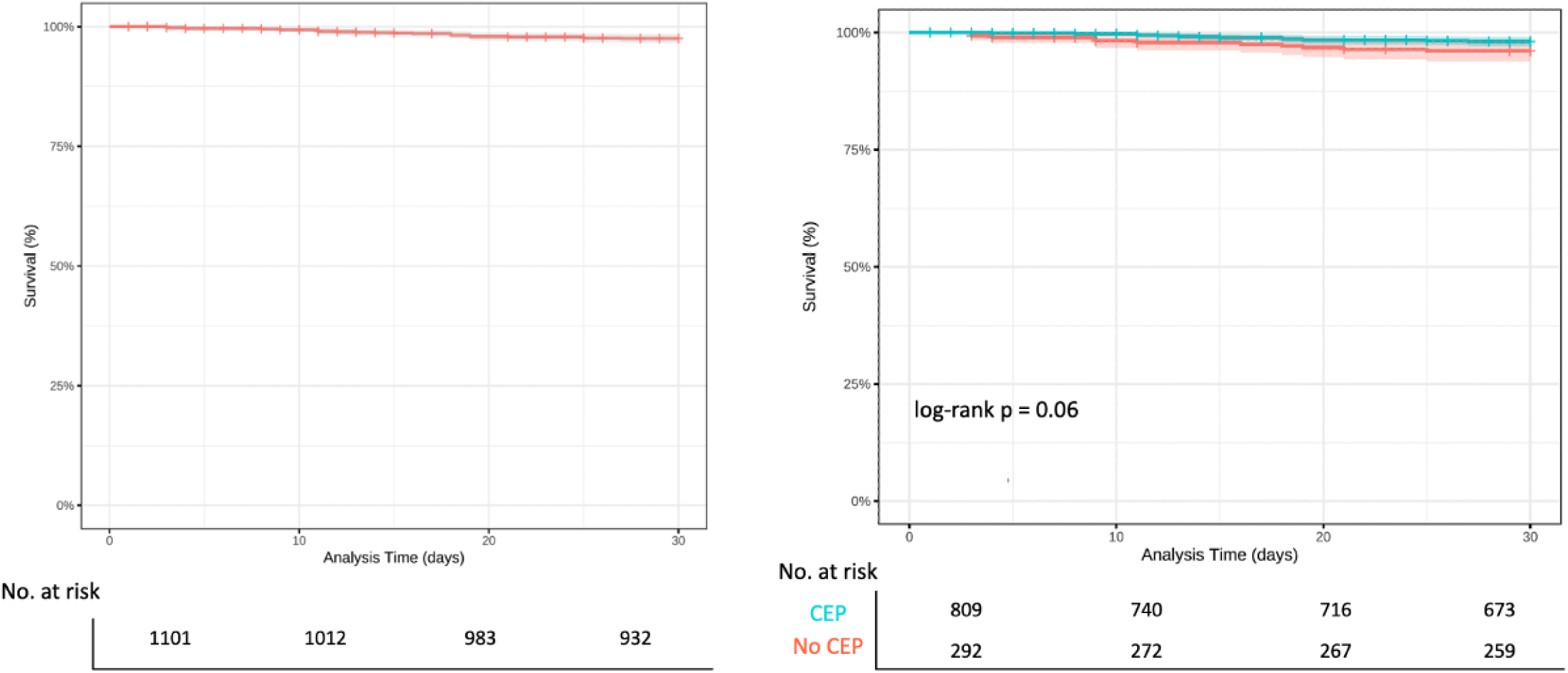
Kaplan-Meier Survival Curve for 30-day mortality. Left: total sample, Right: Blue: transfemoral TAVR with CEP, Red: transfemoral TAVR without CEP. Survival at 30 days follow up does not significantly differ between patients undergoing TAVR with or without neuroprotection devices - log-rank p = 0.06.

Furthermore, the Kaplan-Meier curves of patients treated between 2019 and 2025 revealed a significantly lower 30-day mortality rate in the CEP group (1.9% [0.9% 2.9%] vs. 4.7% [1%; 8.4%] Cox regression: HR 0.38, 95% CI: (0.14-0.98), p = 0.05)) (Supplementary Figure 2). (Supplementary Table 1).

## Discussion

This study provides several key findings: 1) both overall neurological events and ischemic stroke within 3 days after TAVR were significantly reduced in the patient group for whom cerebral embolic protection (CEP) was possible; 2) CEP use was associated with > 65% relative risk reduction of ischemic stroke; 3) CEP had no statistically significant association with TIA and delirium.

Although technological refinements and operator experience have reduced many procedural complications, periprocedural stroke risk remained relatively stable at 2-5% over the last decade, varying by patient risk profile ^11^. Early trials such as PARTNER-1A ^12^ reported higher stroke rates in TAVR compared with SAVR (5.5% vs. 2.4%, p = 0.04). More recent data from PARTNER II and III trials ^13,14^ have shown lower rates (0.6-3%), yet overall stroke incidence has not declined despite expansion of TAVR into younger, lower-risk cohorts ^15^.

Stroke has a multifactorial etiology, embolization of valve-, vessel- or procedure related debris is considered the primary mechanism ^16^. Consistent with prior reports, neurological events in our cohort occurred predominantly within 3 days, with more than half on the procedural day, mirroring findings from Huded et al. ^17^.

### CEP and Neurological Outcomes

Since the introduction of CEP devices in 2012, multiple imaging, histopathologic and clinical studies have been conducted to test the feasibility and safety of neuroprotection devices. In the early feasibility studies, filters captured debris in 80-99% of cases, including thrombus, tissue, and foreign material ^4,18^.

Imaging-dedicated randomized clinical trials (RCTs), such as CLEAN TAVI ^3^, MISTRAL-C ^5^, REFLECT II ^6^ and DEFLECT III ^7^ showed that CEP reduced the number and volume of new MRI lesions and may mitigate neurocognitive decline. With TAVR increasingly implanted in younger patients, the potential long-term consequences of silent brain injuries linked to neurocognitive decline, are of growing relevance ^19,20^. In MISTRAL-C trial neurocognitive deterioration was 7 times more frequent in the unprotected cohort (4% vs. 27%; p = 0.017) ^5^, whereas DEFLECT III trial showed better memory performance, fewer neurological deficits and an increased cognitive recovery with use of TriGuard ^7^. Lesion volume, in particular, has emerged as predictive marker for stroke, with larger total lesion burden > 500 mm^3^ correlating strongly with increased and disabling stroke risk ^21^.

Large RCTs evaluating clinical endpoints, PROTECTED TAVR ^8^ and BHF PROTECT-TAVI ^9^ provide more nuanced results. PROTECTED TAVR showed a non-significant 22% relative reduction in stroke (2.1% and 2.7%), whereas our study observed a larger difference (1.4% [95% CI: 0.6%; 2.2%] vs 4.1% [95% CI: 1.8%; 6.4%]). A post-hoc US-only analysis of PROTECTED TAVR by Makkar et al. ^22^ demonstrated a significant stroke reduction with CEP, suggesting regional differences in case selection, procedural strategies or baseline risk may have influenced results. Importantly, the PROTECTED cohort had a substantially lower baseline risk (mean STS ≈ 3%) than our population (mean STS ≈10%), which may explain the higher stroke rates in unprotected patients in our study. Notably, the PROTECTED trial did demonstrate a reduction in disabling stroke in the CEP group (0.5% vs. 1.3%, p = 0.02). However, the larger BHF PROTECT-TAVI trial, enrolling > 7600 patients, found no difference in overall (2.1% vs. 2.2%) or disabling stroke (1.2% vs. 1.4%). A substudy from BHF PROTECT- TAVI by Kennedy et al. ^23^ also reported no cognitive benefit with CEP, though the absence of systematic neuroimaging limits conclusions, given the high rate of subclinical cognitive decline observed. Both RCTs were ultimately underpowered due to unexpectedly events rates; contemporary estimates suggest that > 22,000 patients would be needed to detect a significant reduction in stroke with CEP at current incidence levels ^24^.

In contrast with RCTs, observational studies consistently report benefit with CEP. Multiple propensity-matched analyses show reductions in early stroke, neurological events and composite endpoints. Seeger et al. ^25^ observed significantly lower 7-day mortality or stroke with CEP (2.1% vs. 6.8%) and stroke rates nearly identical to ours (1.4% vs. 4.6%).

Importantly, we used two different CEP systems (when available on the market) to adapt to patient’s anatomy. This was not possible in company sponsored RCTs and may explain part of the observed difference. While the SENTINEL system was routinely used, patients with unavailable radial access or soft plaques in the truncus were protected by the TRIGUARD system. Thereby, we were able to improve the overall percentage of patients protected, and reduced device related adverse events.

Larger meta-analyses further support CEP effectiveness. Shahid’s analysis of > 400,000 patients reported significantly reduced stroke and disabling stroke with CEP ^26^, while Kheyrbek highlighted a clear difference between RCT and observational data, with benefit confined primarily to real-world cohorts ^27^. Another meta-analysis pooling CLEAN-TAVI, MISTRAL-C, and PROTECTED showed reduced overall and disabling stroke with a notably lower number needed to treat than reported in PROTECTED TAVR ^24^. Our findings align closely with these observational results, reinforcing the signal of reduced early stroke with CEP.

### CEP and Mortality

Periprocedural stroke is a strong determinant of mortality. Although the difference was not statistically significant, our results suggest a trend towards lower 30-day mortality with neuroprotection devices. Prior analyses have shown a >3-fold increase in 30-day mortality after stroke ^28^ consistent with our finding which showed that ischemic stroke within 3 days independently predicted 30-day mortality [HR 7.04, 95% CI: (2.11-23.52), p = 0.002]. While some earlier observational studies reported similar in-hospital and 30-day mortality rates between both groups ^2,29^, two recent meta-analyses suggest that CEP may improve both, neurological events and all-cause mortality ^26,27^.

## Conclusion

In our study CEP was associated with a significant reduction in ischemic stroke occurring within 3 days. Importantly, two different CEP systems were used if available to adapt to patient’s anatomy. Ischemic stroke was a significant predictor of death at 30 days after TAVR. Although CEP use was not associated with a significant reduction in mortality, we observed a trend towards lower 30-day mortality rates in the CEP group.

## Limitations

We conducted a single-center retrospective analysis with inherent limitations. The use of CEP was determined by our cardiac surgery team considering anatomy and technical feasibility. Since we started using CEP in October 2019, earlier patients were operated without, which is a cofounding factor when comparing both groups. Due to the study’s observational nature selection bias and baseline heterogeneity cannot be excluded, why our findings should be interpreted hypothesis generating.

## Data Availability

Data supporting the findings of this study will be provided by the corresponding author upon request.

## Ethical Approval

The present study was approved by the Ethics Committee of the Medical University of Vienna (Approval Number: 1680/2020).

## Informed Consent Statement

The informed consent requirement was waived.

## Non-standard Abbreviations and Acronyms

CEPs: cerebral embolic protection devices
CT: computed tomography
CNS: central nervous system
MRI: magnetic resonance imaging
NeuroARC: Neurologic Academic Research Consortium
NOACs: non-vitamin K oral anticoagulants
RCTs: randomized clinical trials
SAVR: surgical aortic valve replacement
STS: Society of Thoracic Surgeons
TF-TAVR: transfemoral transcatheter aortic valve replacement
TIA: transient ischemic attack
VARC 3: Valve Academic Research Consortium-3

## Supplemental Material

**Supplementary Figure 1.** Kaplan-Meier Survival Curve of 30-day mortality for the subset 2019-2025

**Supplementary Table 1.** Primary and secondary outcomes in the subset 2019-2025

### Central illustration

Impact of Cerebral Embolic Protection Devices on Periprocedural Neurological Outcomes and 30-day Mortality

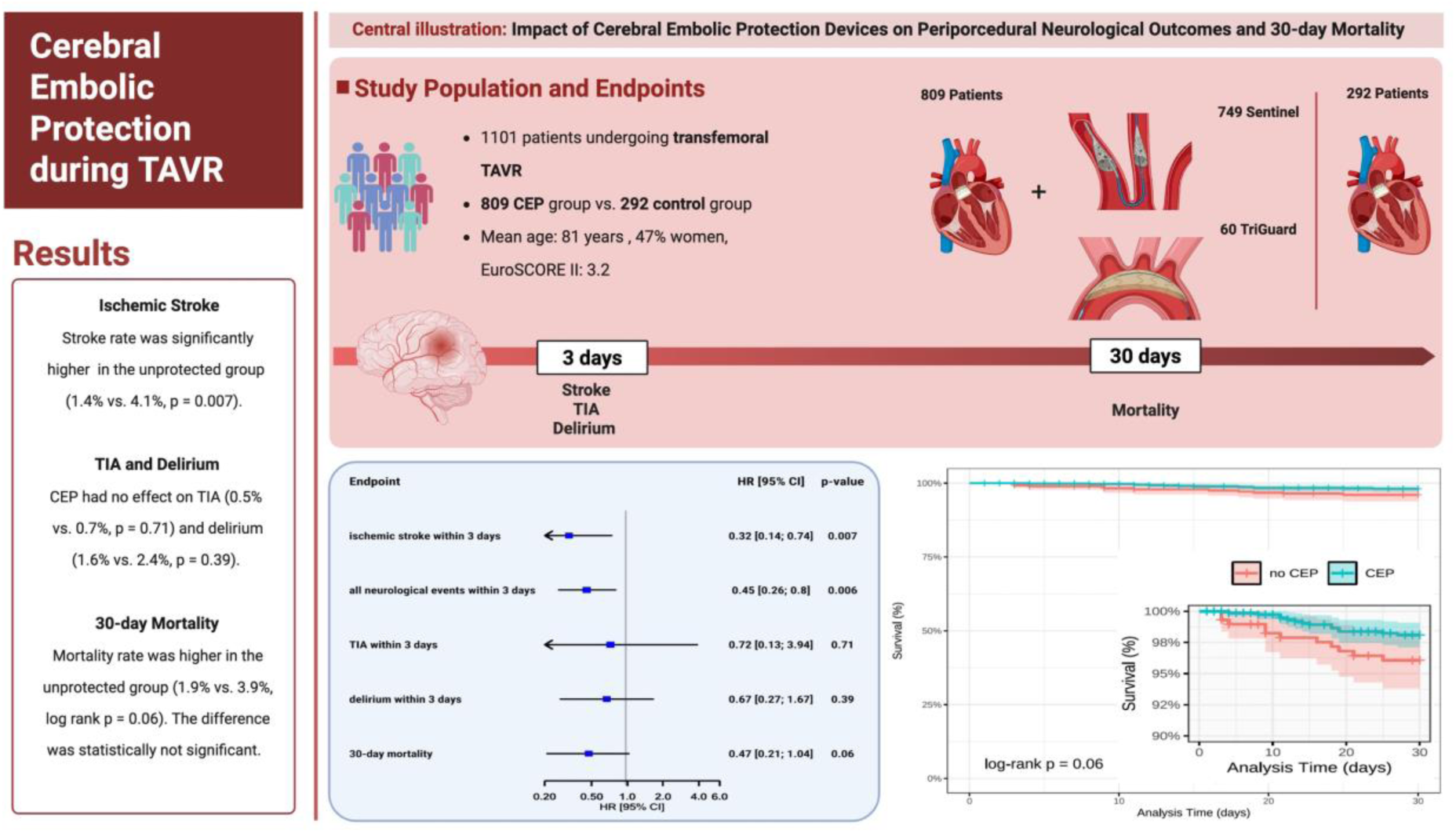

Legend Central Illustration: Overview of Study Population and Outcomes. Created in BioRender. Baysal, F. (2026)

## References

1. Iskander M, Jamil Y, Forrest JK, Madhavan MV, Makkar R, Leon MB, Lansky AJ, Ahmad Y. Cerebral embolic protection in transcatheter aortic valve replacement. Struct Heart. 2023;7:100169. doi: 10.1016/j.shj.2023.100169

2. Shekhar S, Isogai T, Agrawal A, Kaw R, Mahalwar G, Krishnaswamy A, Puri R, Reed G, Mentias A, Kapadia S. Outcomes and predictors of stroke after transcatheter aortic valve replacement in the cerebral protection device era. J Am Heart Assoc. 2024;13:e034298. doi:10.1161/JAHA.124.034298

3. Haussig S, Mangner N, Dwyer MG, Lehmkuhl L, Lucke C, Woitek F, Holzhey DM, Mohr FW, Gutberlet M, Zivadinov R, et al. Effect of a cerebral protection device on brain lesions following transcatheter aortic valve implantation in patients with severe aortic stenosis: the CLEAN-TAVI randomized clinical trial. JAMA. 2016;316:592–601. doi:10.1001/jama.2016.10302

4. Kapadia SR, Kodali S, Makkar R, Mehran R, Lazar RM, Zivadinov R, Dwyer MG, Jilaihawi H, Virmani R, Anwaruddin S, et al. Protection against cerebral embolism during transcatheter aortic valve replacement. J Am Coll Cardiol. 2017;69:367–377. doi:10.1016/j.jacc.2016.10.023

5. Van Mieghem NM, van Gils L, Ahmad H, van Kesteren F, van der Werf HW, Brueren G, Storm M, Lenzen M, Daemen J, van den Heuvel AF, et al. Filter-based cerebral embolic protection with transcatheter aortic valve implantation: the randomised MISTRAL-C trial. EuroIntervention. 2016;12:499–507. doi:10.4244/EIJV12I4A84

6. Nazif TM, Moses J, Sharma R, Dhoble A, Rovin J, Brown D, Horwitz P, Makkar R, Stoler R, Forrest J, et al. Randomized evaluation of TriGuard 3 cerebral embolic protection after transcatheter aortic valve replacement: REFLECT II. J Am Coll Cardiol Intv. 2021;14:515–527. doi:10.1016/j.jcin.2020.11.011

7. Lansky AJ, Schofer J, Tchetche D, Stella P, Pietras CG, Parise H, Abrams K, Forrest JK, Cleman M, Reinöhl J, et al. A prospective randomized evaluation of the TriGuard HDH embolic DEFLECTion device during transcatheter aortic valve implantation: results from the DEFLECT III trial. Eur Heart J. 2015;36:2070–2078. doi:10.1093/eurheartj/ehv191

8. Kapadia SR, Makkar R, Leon M, Abdel-Wahab M, Waggoner T, Massberg S, Rottbauer W, Horr S, Sondergaard L, Karha J, et al. Cerebral embolic protection during transcatheter aortic-valve replacement. N Engl J Med. 2022;387:1253–1263. doi: 10.1056/NEJMoa2204961

9. Kharbanda RK, Kennedy J, Jamal Z, Dodd M, Evans R, Bal KK, Perkins AD, Blackman DJ, Hildick-Smith D, Banning AP, et al. Routine cerebral embolic protection during transcatheter aortic-valve implantation. N Engl J Med. 2025;392:2403–2412. doi:10.1056/NEJMoa2415120

10. Mehta VC, Chandrasekhar SA, Quimby DL, Bhandari A, Mazo V, Glaser AD, Rose DZ, Mohanty BD. Cerebral protection in trans-catheter aortic valve replacement: review and contemporary assessment of randomized trial data. Neurohospitalist. 2024;14:157–165. doi:10.1177/19418744231225680

11. Gasior T. 2024 update on cerebral embolic protection after transcatheter aortic valve replacement. J Clin Med. 2024;13:7256. doi: 10.3390/jcm13237256

12. Smith CR, Leon MB, Mack MJ, Miller DC, Moses JW, Svensson LG, Tuzcu EM, Webb JG, Fontana GP, Makkar RR, et al. Transcatheter versus surgical aortic-valve replacement in high-risk patients. N Engl J Med. 2011;364:2187–2198. doi:10.1056/NEJMoa1103510

13. Vahanian A, Urena M, Walther T, Treede H, Wendler O, Lefèvre T, Spence MS, Redwood SR, Kahlert P, Rodés-Cabau J, et al. Thirty-day outcomes in patients at intermediate risk for surgery from the SAPIEN 3 European approval trial. EuroIntervention. 2016;12:e235–e243. doi:10.4244/eijv12i2a37

14. Mack MJ, Leon MB, Thourani VH, Makkar R, Kodali SK, Russo M, Kapadia SR, Malaisrie SC, Cohen DJ, Pibarot P, et al. Transcatheter aortic-valve replacement with a balloon-expandable valve in low-risk patients. N Engl J Med. 2019;380:1695–1705. doi:10.1056/NEJMoa1814052

15. Reddy P, Merdler I, Ben-Dor I, Satler LF, Rogers T, Waksman R. Cerebrovascular events after transcatheter aortic valve implantation. EuroIntervention. 2024;20:e793–e805. doi:10.4244/EIJ-D-23-01087

16. Nikas DN, Lakkas L, Nikopoulos S, Tsamis K, Sakellariou X, Florentin M, Papanagiotou P, Naka KK, Ntaios G, Michalis L. Stroke in transcatheter aortic valve implantation (TAVI): A comprehensive review. J Clin Med. 2025;14. doi:10.3390/jcm14196754

17. Huded CP, Tuzcu EM, Krishnaswamy A, Mick SL, Kleiman NS, Svensson LG, Carroll J, Thourani VH, Kirtane AJ, Manandhar P, et al. Association between transcatheter aortic valve replacement and early postprocedural stroke. JAMA. 2019;321:2306–2315. doi:10.1001/jama.2019.7525

18. Van Mieghem NM, Schipper ME, Ladich E, Faqiri E, van der Boon R, Randjgari A, Schultz C, Moelker A, van Geuns RJ, Otsuka F, et al. Histopathology of embolic debris captured during transcatheter aortic valve replacement. Circulation. 2013;127:2194–2201. doi:10.1161/CIRCULATIONAHA.112.001091

19. Jimenez Diaz VA, Kapadia SR, Linke A, Mylotte D, Lansky AJ, Grube E, Settergren M, Puri R. Cerebral embolic protection during transcatheter heart interventions. EuroIntervention. 2023;19:549–570. doi:10.4244/EIJ-D-23-00166

20. Woldendorp K, Indja B, Bannon PG, Fanning JP, Plunkett BT, Grieve SM. Silent brain infarcts and early cognitive outcomes after transcatheter aortic valve implantation: a systematic review and meta-analysis. Eur Heart J. 2021;42:1004–1015. doi:10.1093/eurheartj/ehab002

21. Lansky AJ, Grubman D, Dwyer MG III, Zivadinov R, Parise H, Moses JW, Shah T, Pietras C, Tirziu D, Gambone L, et al. Clinical significance of diffusion-weighted brain MRI lesions after TAVR: results of a patient-level pooled analysis. J Am Coll Cardiol. 2024;84:712–722. doi:10.1016/j.jacc.2024.05.055

22. Makkar RR, Gupta A, Waggoner TE, Horr S, Karha J, Satler L, Stoler RC, Alvarez J, Sakhuja R, MacDonald L, et al. Cerebral embolic protection by geographic region: a post hoc analysis of the PROTECTED TAVR randomized clinical trial. JAMA Cardiol. 2025;10:17–24. doi:10.1001/jamacardio.2024.4278

23. Kennedy J, Blackman DJ, Dodd M, Poggesi A, Read L, Jamal Z, Evans R, Clayton T, Kharbanda RK, Hildick-Smith D. Impact of cerebral embolic protection on cognitive function after transcatheter aortic valve implantation: data from the BHF PROTECT-TAVI randomized trial. Circulation. 2025;152:1268–1278. doi:10.1161/CIRCULATIONAHA.125.076761

24. Wolfrum M, Handerer IJ, Moccetti F, Schmeisser A, Braun-Dullaeus RC, Toggweiler S. Cerebral embolic protection during transcatheter aortic valve replacement: a systematic review and meta-analysis of propensity score matched and randomized controlled trials using the Sentinel cerebral embolic protection device. BMC Cardiovasc Disord. 2023;23:306. doi:10.1186/s12872-023-03338-0

25. Seeger J, Gonska B, Otto M, Rottbauer W, Wöhrle J. Cerebral embolic protection during transcatheter aortic valve replacement significantly reduces death and stroke compared with unprotected procedures. J Am Coll Cardiol Intv. 2017;10:2297–2303. doi:10.1016/j.jcin.2017.06.037

26. Shahid S, Kasbati M, Adawi SO, Ishtiaq S, Osama M, Shah SMA, Saifullah M, Ali S, Ahmed R, Ch IA, et al. Efficacy of cerebral embolic protection device in transcatheter aortic valve replacement: a systematic review and meta-analysis. Catheter Cardiovasc Interv. 2025;106:2996–3007. doi:10.1002/ccd.70146

27. Kheyrbek M, Alsabti S, Niroula S, Ahmad E, Choucair M, Bhatia U, Wernette A, Chhabra K, Strubchevska K, Hanson I, et al. Safety and efficacy of cerebral embolic protection systems in transcatheter aortic valve replacement: a systematic review and meta-analysis. Expert Rev Cardiovasc Ther. 2024:1–8. doi:10.1080/14779072.2024.2445256

28. Eggebrecht H, Schmermund A, Voigtlander T, Kahlert P, Erbel R, Mehta RH. Risk of stroke after transcatheter aortic valve implantation (TAVI): a meta-analysis of 10,037 published patients. EuroIntervention. 2012;8:129–138. doi:10.4244/EIJV8I1A20

29. Kroon HG, van der Werf HW, Hoeks SE, van Gils L, van den Berge FR, El Faquir N, Rahhab Z, Daemen J, Poelman J, Schurer RAJ, et al. Early clinical impact of cerebral embolic protection in patients undergoing transcatheter aortic valve replacement. Circ Cardiovasc Interv. 2019;12:e007605. doi:10.1161/CIRCINTERVENTIONS.118.007605

